# Assessing microstructural damage in paediatric aortic coarctation tissue during benchtop balloon angioplasty and stenting

**DOI:** 10.1101/2024.10.22.24315705

**Authors:** N. Linnane, R. D Johnston, D. Kenny, C. Lally

## Abstract

Endovascular stenting of native aortic coarctation has become standard treatment for children and young adults due to the shorter hospital stay and recovery time, lower procedural risk and less invasive nature. However, despite these advantages, neonatal coarctation remains a surgically treated lesion, owing to the challenges of vessel growth and stent design. Plain old balloon angioplasty (POBA) of neonatal coarctation has been used as a treatment modality in the past but concern around vessel damage and aneurysm formation limited its widespread adoption. To-date, stenting has often been preferred over POBA treatment to minimise tissue damage but very little evidence has been gathered to establish the degree of tissue damage during intravascular procedures in neonatal coarctation tissue.

The aim of this study was to perform balloon angioplasty and stenting on coarctation the aorta samples harvested at the time of surgery from neonatal patients and visualise the deformation under ultrasound guidance. From this, the novel collagen hybridizing peptide (CHP) was used to assess the level of collage denaturation and microstructural damage this tissue experienced when undergoing these procedures. The results demonstrate that balloon angioplasty singularly is not a viable treatment option at maintaining the lumen diameter unless higher levels of stretch are achieved to permanently deform the tissue. Five out of six neonatal coarctation samples were successfully stented with no stent failure, proving that stent insertion is a viable option to achieve the desired lumen diameter. In addition, for both procedures, increased collagen damage was observed with higher tissue strain. Overall, this work would suggest stent placement as the best option to achieve reliable lumen gain and to limit the level of the level of collagen damage when stretching the tissue to achieve the required lumen diameter.

## Introduction

Coarctation of the aorta (CoA) is reported to have an incidence of 0.3 to 0.6 in 1000 live births, comprising for 5% - 8% of all congenital heart diseases [1,2]. Technically, it is defined as a narrowing of the aorta, typically located at the insertion of the ductus arteriosus just distal to the left subclavian artery, although it can be located either proximally or distally to the ductus [3–5]. The narrowing is often discrete but, in some cases, can be a long segment stenosis and/or tortuous [3,5]. A discrete coarctation can occur in isolation, however it is commonly associated with other congenital heart defects, including bicuspid aortic valve (60%), aortic arch hypoplasia and other arch anomalies (18%), ventricular septal defect (13%), mitral valve abnormalities (8%) and sub-aortic stenosis (6%) [3]. Historically, CoA was treated with surgical excision and anastomosis to remove the obstruction. However, in recent times, endovascular treatment options including balloon angioplasty and stent implantation have been described [2,6,7]. The reason for this alteration in procedure is due to the reported lower procedural risk and shorter recover times associated with endovascular based approaches when compared to surgery [5,8,9].

Initial reporting into evaluating the local tissue response to percutaneous therapy was by Lock et al (1982), who harvested seven coarctation specimens at the time of surgery and performed in vitro balloon angioplasty within two hours of harvest. They demonstrated intima and media tears but relief of the obstruction, paving the way for safety and efficacy studies to be undertaken [10]. Shaddy et al (1993) performed the first randomised trial of 36 patients comparing balloon angioplasty (20 patients) to surgical treatment (16 patients) for native coarctation of the aorta. It was observed that there was a reduction in the pressure gradient with few complications noted. Furthermore, there was a higher rate of restenosis in the balloon angioplasty group, yet this did not reach statistical significance [8]. The complications observed such as higher aneurysm formation in the balloon angioplasty group could be explained by Lock et al (1982) observations, whereby intima and media damage were a necessary compromise for successful pressure gradient reduction [10,11]. Interestingly, Forbes et al (2007) reported on a large series of 578 stenting procedures for the treatment of aortic coarctation, demonstrating excellent outcomes, however aortic wall injury such as aneurysm or dissection could occur [12]. This observation was supported by long term data from the Coarctation of the Aorta Stent Trial (COAST) demonstrating aneurysm formation as late as 3 years post procedure [23]. Forbes et al (2007) through the Congenital Cardiovascular Interventional Study Consortium (CCISC) report that higher morbidity and mortality is observed in older patients, likely due to reduced aortic compliance [12]. Furthermore, the CCISC report followed on from this with a prospective study by Holzer et al (2010) showing increased wall injury with balloon angioplasty compared to surgery or stenting [13]. There are multiple other studies which have demonstrated the safety and efficacy of stenting in coarctation of the aorta and this is becoming standard care in many institutions [14–21]. The issue is that stents used in treatment of CoA are generally “off-label”, with biliary stents being the most commonly used, leaving less than optimum outcomes [22–24]. However, it is important to note that there are some promising recent studies looking at developing stents specifically for the treatment of coarctation of the aorta in infants [25,26].

Although stenting is improving patient outcomes, due to excessive expansion, the tissue is being damaged during balloon angioplasty and stenting, leading to possible aneurysm development or aortic dissection [12,27–29]. A recent study by Cornelissen et al (2023), also observed that the magnitude of stent indentation is positively correlated to level of vessel damage, with subsequent neointimal growth observed in histology [30]. To-date however, there has been no study to quantify the level of microstructural damage observed in specifically neonatal aortic coarctation tissue during balloon angioplasty and stenting.

To assess microstructural damage, collagen hybridizing peptide (CHP) has been used in a range of tissues to date [31–40]. CHP is known to be a synthetic peptide with the collagen mimicking glycine-proline-hydroxyproline repeating sequence and has a strong propensity for forming the triple helical structure of collagen [39–41]. When the collagen is damaged / denatured, the triple helix unravels allowing for the CHP to bind to reform the triple helix structure. CHP can also be fluorescently labelled which allows for damaged collagen to be localized with the tissue and ultimately quantified [39–41]. Converse et al (2018) was the first to show that CHP is sensitive to stretch induced collagen damage in cerebral arteries, with the highest fluorescence observed in the medial layer, the layer which is known to be the load dominant layer, indicating highest level of damage [35]. Zitnay et al (2020) interestingly showed collagen denaturation increased with increasing cycles of fatigue loading in tendons [33] and Anderl et al (2023) showed that strain induced collagen damage in cerebral vessels is rate dependent, with significantly less collagen denaturation at high rates in aligned fibres [37]. A more recent study by Smith et al (2023) showed that collagen molecular damage is indicative of early atherosclerotic development, with higher levels of CHP observed in ApoE mice on a high fat diet over time [34]. The observations in these studies are interesting as balloon angioplasty and stenting can impose high supraphysiolgical levels of stress and strain within the tissue [42–45]. Furthermore, it is known through imaging evaluation that there is significant collagen fibre remodelling observed in the tissue during stent strut indentation [46,47], suggesting significant microstructural change. However, no study to date, has assessed the level of microstructural damage that could occur within underdeveloped neonatal aortic coarctation tissue when undergoing balloon angioplasty and stenting.

In this study, balloon angioplasty and stenting of neonatal aortic coarctations are performed under ultrasound guidance in an in-house benchtop setup to see the best treatment method in restoring luminal diameter. For the first time, ballooned and stented neonatal coarctation samples are then analysed further using the novel collagen hybridizing peptide (CHP) to assess the level of microstructural damage induced during these treatment procedures.

## Materials & Methods

### In-vivo CoA vessel imaging

To determine the optimum angioplasty balloon and stent sizing, initially, echocardiographic images of neonatal aortic coarctation patients were taken to establish the size of the isthmus and descending aorta. Sandovel et al (2020) recommended that the balloon for angioplasty and stenting should be 2-3 times larger than the isthmus, but not larger than the descending aorta [48]. As shown in figure 1A and 1B, the suprasternal view was used to isolate the isthmus and the long axis subcostal view was used to visualize the descending aorta, whereby geometric measurements were extracted (yellow lines) and recorded, see Tables 1 and 2. The balloon size was recorded before balloon angioplasty and stenting and the balloon to isthmus ratio was calculated for grouped datasets in our analysis.

**Figure 1.**
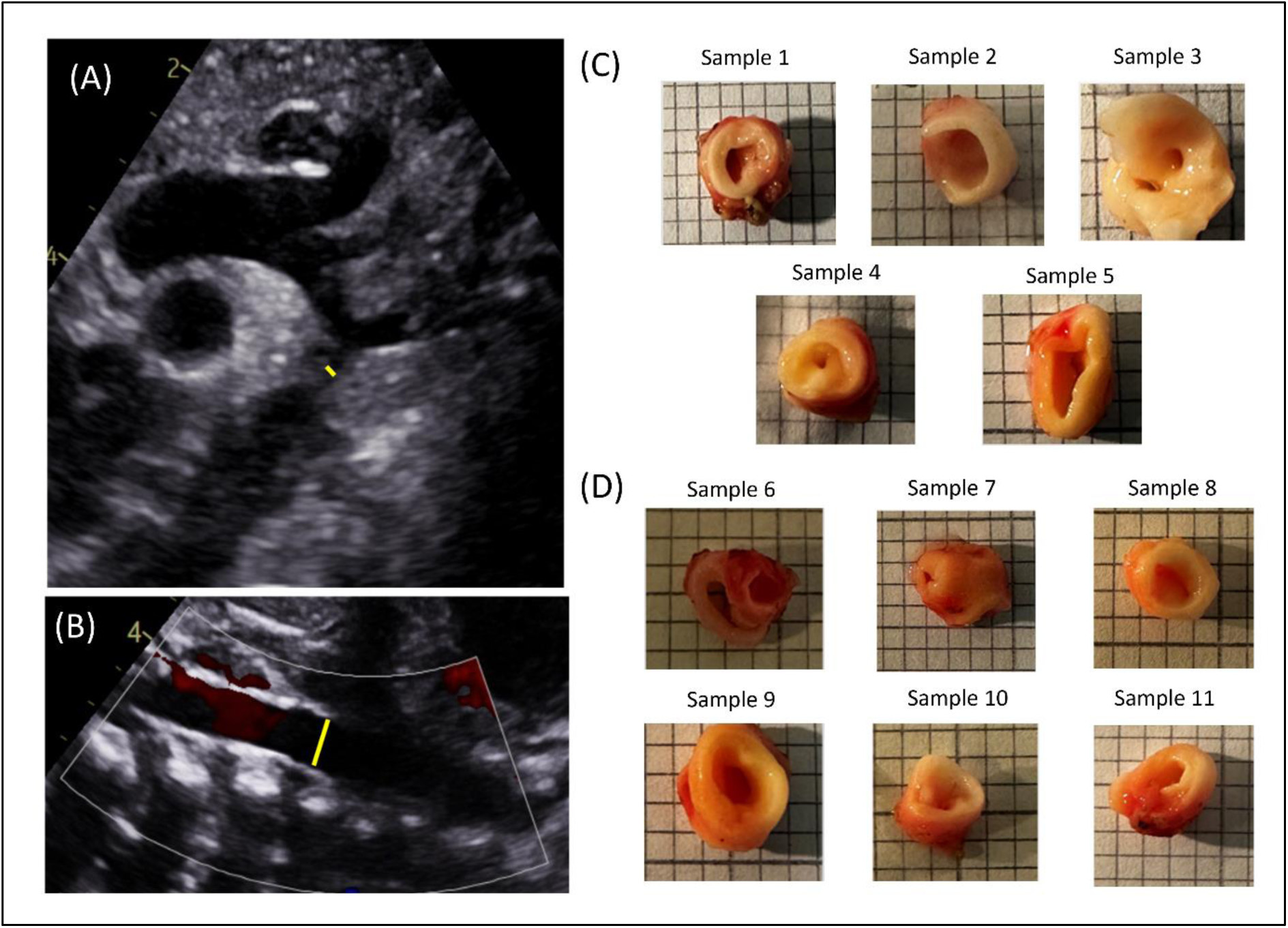
Echocardiographic images used to take measurements prior to benchtop testing, (A) Suprasternal view where isthmus measurement (yellow line) was taken, (B) Long axis subcostal view where descending aorta measurement at diaphragm (yellow line) was taken, (C) Photographic images of samples 1-5 used for neonatal aortic coarctation balloon angioplasty experiment, (D) Photographic images of samples 6 – 11 used in neonatal aortic coarctation stenting experiment.

**Table 1:** Isthmus and descending aorta measurements taken from echocardiography images of samples 1-5 used for balloon angioplasty experiment. Balloon to isthmus ratio calculated considering balloon size used for expansion of samples.

| Sample Number | Isthmus measurement (mm) | Balloon size (mm) | Descending aorta measurement (mm) | Ratio (Balloon / Isthmus) |
| --- | --- | --- | --- | --- |
| 1 | 1.5 | 4.0 | 5.7 | 2.67 |
| 2 | 2.8 | 4.0 | 4.1 | 1.43 |
| 3 | 2.2 | 5.0 | 4.8 | 2.27 |
| 4 | 3.8 | 5.0 | 4.5 | 1.32 |
| 5 | 3.9 | 6.0 | 6.7 | 1.54 |

**Table 2:** Isthmus and descending aorta measurements taken from echocardiography images of samples 6 - 11 used for aortic coarctation stenting experiment. Balloon to isthmus ratio calculated considering balloon size used for expansion of samples.

| **Sample Number** | **Isthmus measurement (mm)** | **Balloon size (mm)** | **Descending aorta measurement (mm)** | **Ratio (Balloon / Isthmus)** |
| --- | --- | --- | --- | --- |
| **6** | 1.9 | 6.0 | 4.9 | 3.16 |
| **7** | 2.9 | 6.0 | 6.1 | 2.07 |
| **8** | 2.1 | 6.0 | 5.1 | 2.86 |
| **9** | 2.2 | 6.0 | 6.1 | 2.73 |
| **10** | 1.9 | 6.0 | 4.5 | 3.16 |
| **11** | 1.3 | 6.0 | 7.1 | 4.62 |

### Tissue specimen collection and preparation for testing

Neonatal aortic coarctation samples were obtained from 11 patients at Children’s Health Ireland at Crumlin, Ireland. Patients were previously identified as having a coarctation (CoA) by the treating medical team, separate to the research team and a decision made to proceed to surgery. Using previously established protocols for atherosclerotic plaque tissue [49], on the day of surgery, samples were collected from theatre upon their immediate removal and stored in phosphate buffered saline (PBS) and stored for transport at room temperature. Samples were initially photographed, and subsequently stored into tissue freezing medium (TFM), comprising of RPM-60 Media, 1.8M DMSO and 0.1M sucrose and placed into a Mr. Frosty cryosystem containing 2-proponol and cryopreserved at −80°C within 6 hours of harvesting. After a period of 12 hours, the samples were removed from the Mr Frosty cryosystem and left at −80°C until the day of stent implantation. It has been established in the literature that this cryopreservation process has a non-significant effect on the tissues’ mechanical properties [49,50]. On the day of testing, the samples were defrosted in PBS at ambient temperature, ready for placement in our benchtop setup. A small section was excised to act as a control for the level of CHP fluorescence in aortic coarctation tissue. 5 samples were assigned to undergo balloon angioplasty, see figure 1C, and 6 samples were assigned to undergo stenting with cobalt chromium Renata minima stents, see figure 1D.

### Bench-top CoA balloon angioplasty and stenting

For our experiments, a custom-made water bath was designed in Solidworks (Dassault Systèmes Simulia Corp., Rhode Island, USA), see figure 2A, and fabricated in polylactic acid (PLA) filament using a Prusa i3 MK3S+ 3D printer (Prusa Research, Prague, Czechia). The bath was designed to ensure it could accommodate a 12L4 linear array ultrasound probe from a SIEMENS ACUSON S2000™ ultrasound system. To ensure a watertight system, 100% infill was used during printing with the outside of the water bath coated in epoxy resin to ensure no leakage of the PBS during testing. To allow for the introduction of the 0.014” Scion blue coronary wire (Asahi Intec, Japan) and balloons, cable glands were attached on the end of the bath as shown in figure 2B. After the relevant apparatus were introduced for the test, these cable glands were tightened along with a Tuohy-Borst, to ensure suspension of the sample on the coronary wire and allowing for the Encore inflator device (Boston Scientific, Galway, Ireland) to inflate the balloon whilst ensuring no leakages occurred. In our balloon angioplasty experiments, a 4mm, 5mm and 6mm diameter, 30mm length Tyshak 2 balloons (Numed, Hopkinton, NY, USA) were used. For the stenting experiments, the Minima stents (Renata Medical, California, USA) were mounted on a 6 x 30 mm sterling balloon (Boston Scientific, Galway, Ireland). After filling of the water bath with PBS, the 12L4 ultrasound probe was positioned at the top of the bath to obtain cross sectional views of the aortic coarctation sample with the probe set at frequency of 9 MHz, as shown in figure 2C. Continuous video feedback was obtained during the ultrasound acquisition, allowing for imaging of the aortic coarctation in the initial state and after balloon angioplasty and stenting, respectively, see figure 2D.

**Figure 2:**
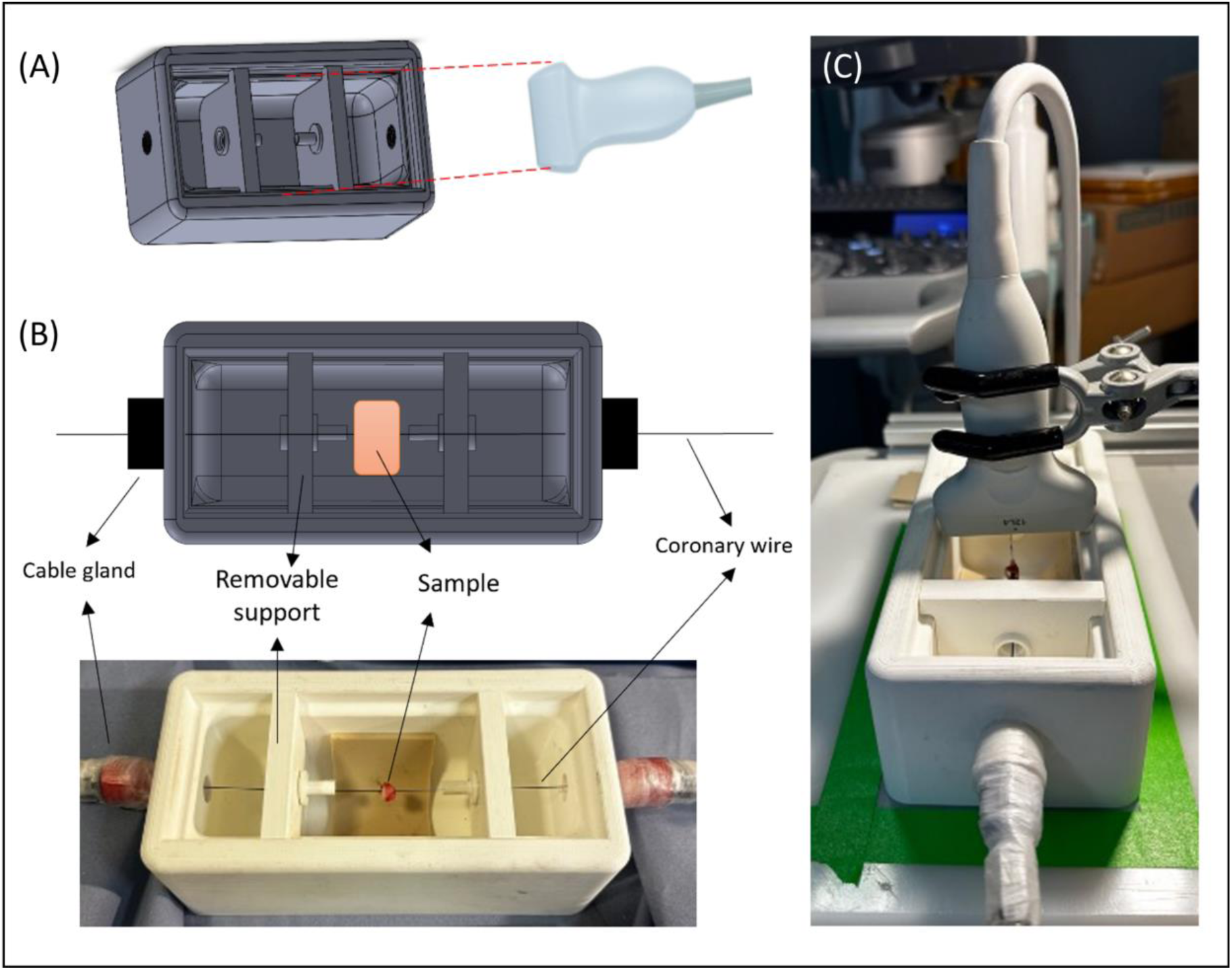
(A) Custom made 3D printed water bath design compatible with ultrasound 12L4 probe (B) Plan view schematic and photograph of the experimental setup showing neonatal aortic coarctation sample suspended on coronary wire before PBS fill (C) Final setup with ultrasound 12L4 probe in position over the neonatal aortic coarctation sample.

### Histological analysis

Balloon expanded and stented neonatal aortic coarctation samples were fixed in 10% formalin for a period of 24 hours. On the day of processing, histology using specific tissue stains, namely Haematoxylin and Eosin (H&E), Picrosirius red (PSR) with polarized light microscopy (PLM) and Verhoeff’s were performed to visualize the cellular content, collagen, and elastin content, respectively. Once fixed, samples were then dehydrated (Leica TP1020, Semi-enclosed benchtop tissue processor, Germany) and embedded in paraffin wax blocks. Following this, 7 µm cross sections were cut from the blocks using C35 microtome blades and subsequently floated on distilled water at 37 °C before mounting on glass slides. The slides were left to dry for a period and staining was then performed using a Leica Autostainer (Leica ST5010, Autostainer XL, Germany), that also performed deparaffinization and rehydration of the samples. Once stained, slides were then cover slipped using a coverslipper (Leica CV5030, Automated Coverslipper Machine, Germany) in which DPX mountant was used as the mounting agent. Brightfield imaging of all stained samples was performed on an Aperio CS2 microscope with ImageScope software V12.3 (Leica Biosystems Imaging, Inc., Vista, California). Polarized light microscopy (PLM) was also performed on the picrosirius red (PSR) stained samples using an Olympus BX41 microscope with Ocular V2.0 software (Teledyne Photometrics, Tuscon, Arizona). Two PLM images were taken for each slice images, with 45° to each other, with the exposure time kept consistent across all samples [49,51].

### Collagen Hybridizing Peptide (CHP)

Collagen hybridizing peptide (CHP) was procured from 3-Helix for the assessment of microstructural damage within the neonatal aortic coarctation under balloon angioplasty and stenting. F-CHP, the type used in this study incorporates a fluorescin component which allows for fluorescence to occur when the CHP binds with the collagen and is subsequently excited. To setup samples for F-CHP staining after testing, the samples are fixed using 10% formalin for a period of 24 hour. Once fixed, the tissue was subsequently processed using standard histological approaches, whereby the sample underwent dehydration (Leica TP1020, Semi-enclosed benchtop tissue processor, Germany) and was embedded in paraffin wax blocks. Following this, 7µm cross sections were cut from paraffin blocks using C35 microtome blades and floated in distilled water at 37 °C before mounting on glass slides. Before F-CHP staining, the slides were rehydrated using a standard protocol of xylene to 100% alcohol to water. F-CHP was diluted in 1ml of PBS and centrifuged making a 100 µM stock solution, which was diluted further to a concentration of 20 µM as recommended for staining slides [31,52] On the day of staining, the F-CHP was heated to 80 °C for 5 mins, cooled rapidly to room temperature and then administered onto samples on glass slides. Stained slides were kept at 4 °C for 24 hours for sufficient binding to occur. For cover slipping, residual F-CHP was rinsed off the samples with PBS and coverslip was applied with Mowiol mounting agent containing DABCO (1,4 diazabicyclo [2.2.2] octane). Once cover slipped, fluorescent imaging of F-CHP stained aortic coarctation samples was performed in a dark room using an Olympus BX51 upright microscope (Olympus Lifescience Solutions). For F-CHP, an excitation wavelength of 495 nm is used with an emission wavelength of 512 nm. Importantly, the exposure time for the images were kept constant for the fluorescent level across samples to be comparable. For the ballooned and stented neonatal tissue, all samples had an exposure of 909.1 ms during imaging. For localized assessment of the fluorescent intensities observed in our F-CHP-stained samples, the software ImageJ [53] was used to extract the mean intensities from the control, ballooned and stented samples respectively.

### Statistical analysis

Statistical analyses were performed using GraphPad Prism 10 (GraphPad Software LLC). Normally distributed data was analysed with unpaired t-test. Non-normally distributed data was analysed using Mann-Whitney test.

## Results

### Balloon angioplasty of neonatal aortic coarctation samples

Samples 1 and 2 were inflated to 4mm and once deflated there was clear recoil of the tissue, enclosing the lumen diameter and no obvious deformation in the images once the balloon was deflated, as shown by figure 3A. Similar results were observed for samples 3 and 4 that were inflated to 5mm diameter, with full recoil occurring after balloon is deflated, see figure 3B. Sample 5 shows that after being inflated to 6mm, there is some level of luminal opening after balloon deflation, see figure 3C. Geometric measurements support visual observations, with little change observed in 4mm expanded samples (2.1mm to 2.4mm and 0.6mm to 0.8mm) and 5mm expanded samples (0.6mm to 0.7mm and 1.1mm to 1.2mm), with the 6mm expanded sample showing a larger lumen diameter then the original diameter (4.7mm to 6.7mm). Overall, these observations would suggest that significant levels of vessel deformation / stretch are required after balloon angioplasty to maintain lumen gain.

**Figure 3:**
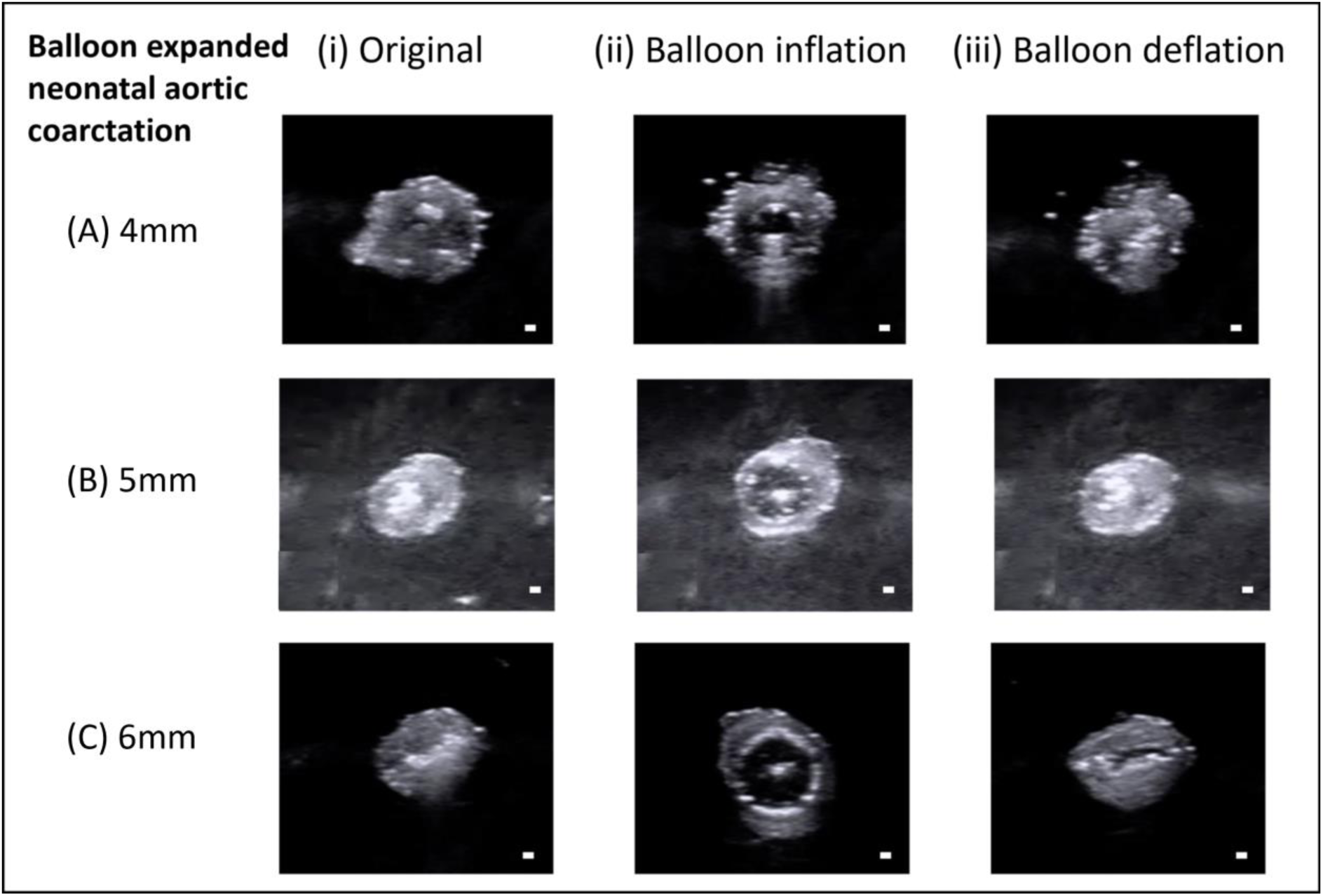
Ultrasound images of the balloon angioplasty experiment with balloon expanded to (A) 4mm, (B) 5mm and (C) 6mm, respectively, depending on the isthmus to descending aorta ratio. Balloon angioplasty experiment images shown represent (i) Original sample (ii) Balloon inflation showing clear opening of the tissue (iii) Balloon deflation showing recoil of the tissue, showing a collapse of the lumen diameter except for the 6atm case, where significant permanent change is observed. Scale bar = 2mm

**Table 3:** Ballooned neonatal samples measurements of lumen diameter before and after ballooning.

| Sample Number | Diameter before<br>ballooning (mm) | Diameter after<br>ballooning (mm) | Percent expansion<br>(%) |
| --- | --- | --- | --- |
| 1 | 2.1 | 2.4 | 14.3 |
| 2 | 0.6 | 0.8 | 33.3 |
| 3 | 0.6 | 0.7 | 16.7 |
| 4 | 1.1 | 1.2 | 9.1 |
| 5 | 4.7 | 6.7 | 42.6 |

### Deployment of stent within neonatal aortic coarctation samples

In the stenting experiment, the Minima stents were successfully deployed into 5 out of 6 neonatal aortic coarctation samples, with sample 10 rupturing during the stent inflation process and not reaching the final 6mm diameter. As shown in figure 4, the result of the 5 successfully stented samples looked similar to the balloon angioplasty experiment, except with full opening of the neonatal aortic coarctation after balloon deflation, and little or no tissue recoil. During the entire procedure, the stent can be visualized under ultrasound imaging with clear stuts being visualized during balloon expansion and deflation. The Renata minima stent performed exactly as expected, with no failure of the stents during this procedure.

**Figure 4:**
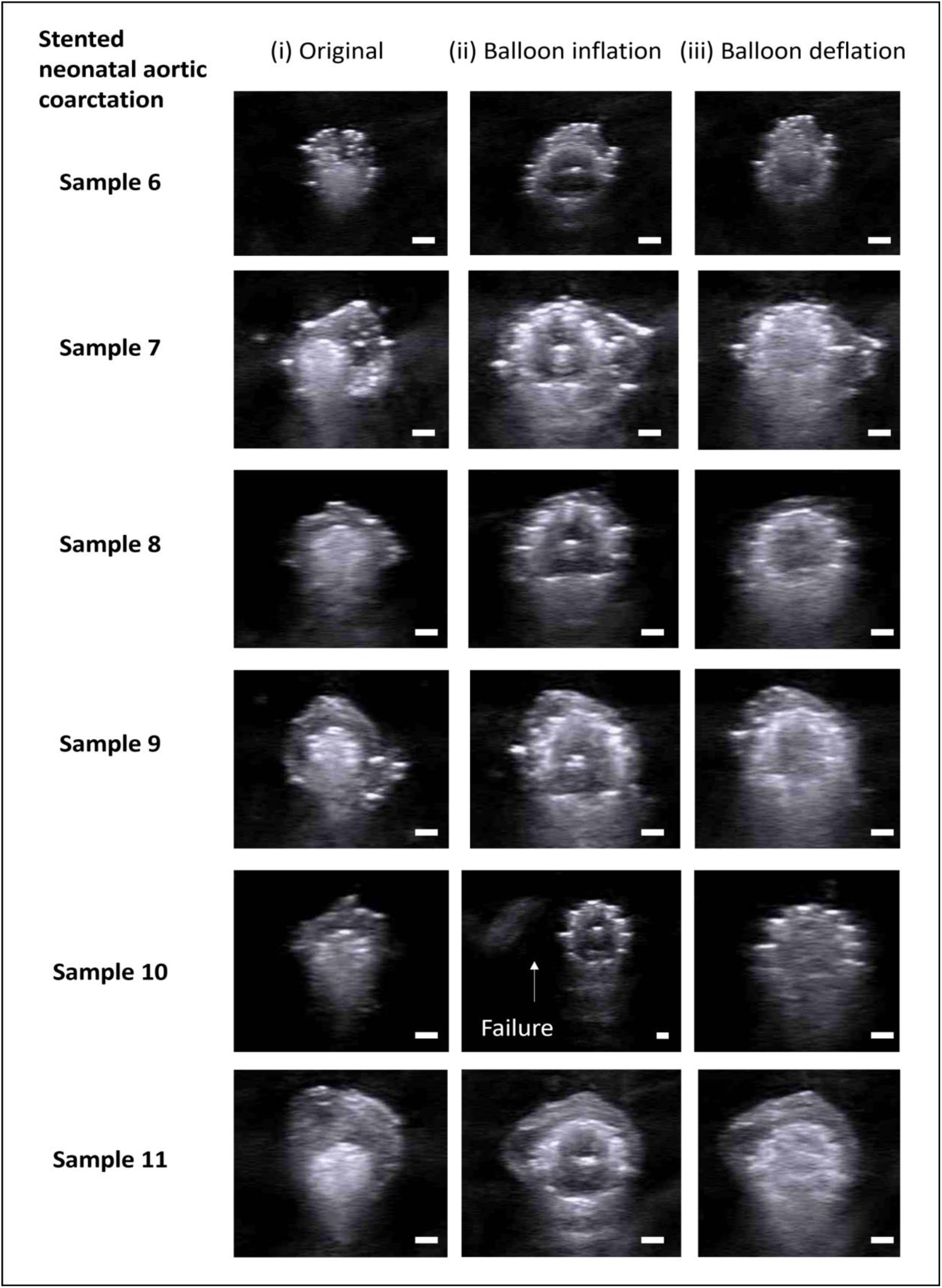
Ultrasound image of stenting procedure of neonatal aortic coarctation using Renata minima stent expanded to 6mm (i) Original (ii) Balloon inflation showing clear opening of the tissue with visible stent struts (iii) Balloon deflation and stent maintaining its diameter. Scale bar = 2mm

### Histological observations of ballooned and stented aortic coarctation samples

Our histological observations confirm the results seen in Johnston & Linnane et al (2024) [Johnston &Linnane 2024] showing neonatal aortic coarctation tissue to be highly heterogenous with collagen and elastin rich areas typical of aortic regions and an absence of structured collagen and elastin in ductal tissue regions, as shown in figure 5A and 5B. An interesting observation can be seen in stented sample 10 which failed during balloon expansion. It is noted histologically that there is ductal tissue propogating through the entire thickness of the sample and due to its low-load bearing capacity, failure is observed.

**Figure 5:**
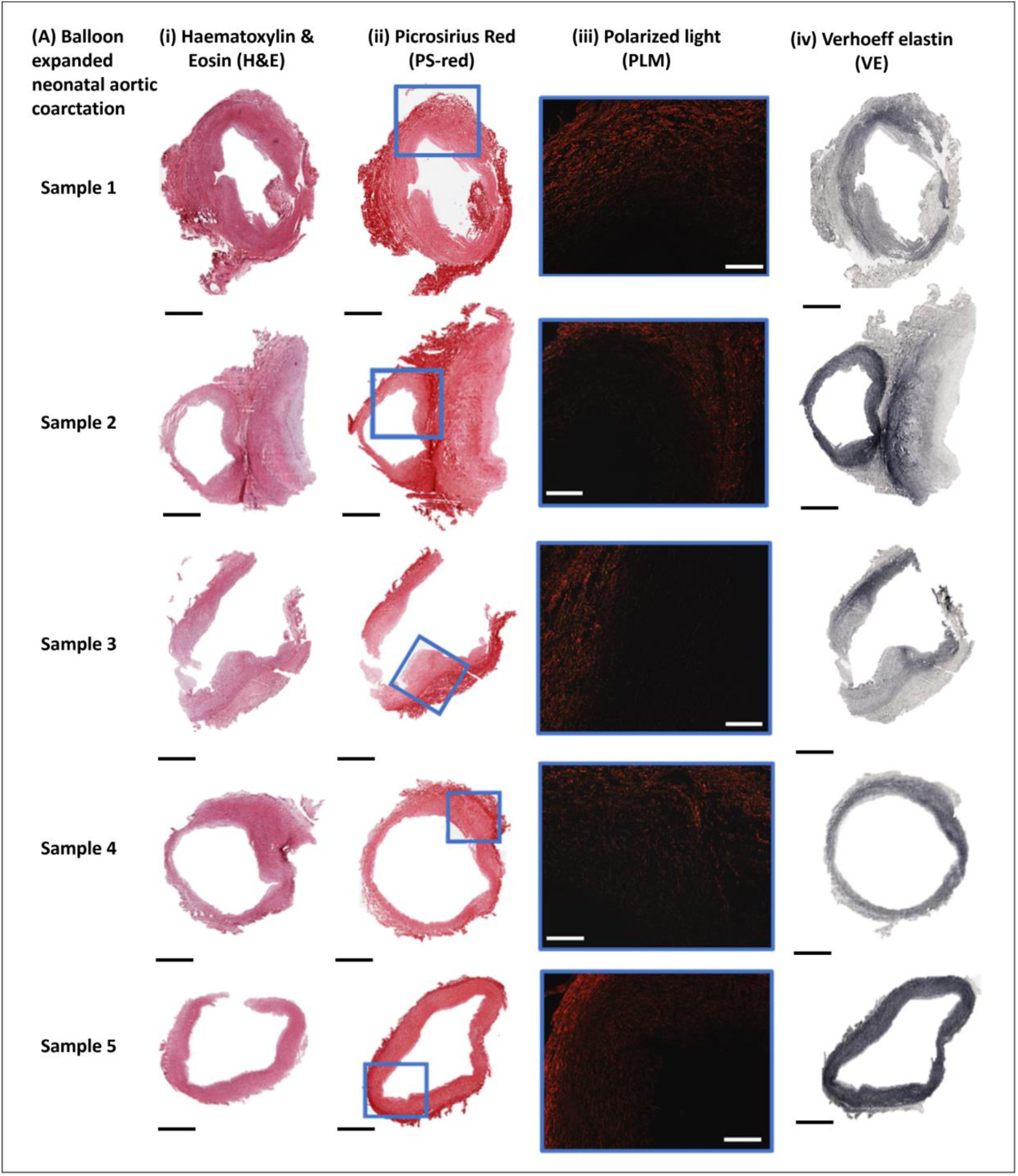
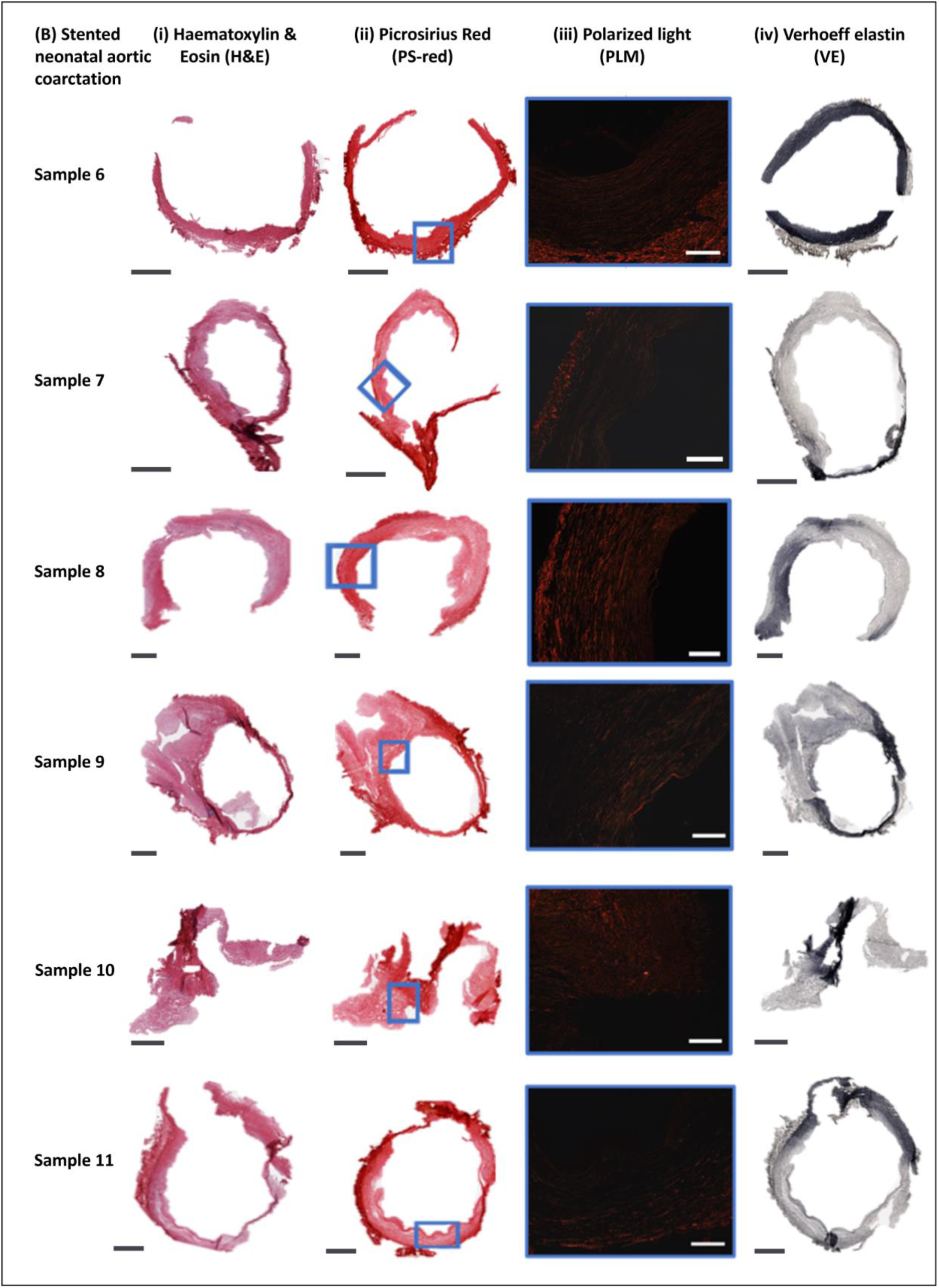
Histological assessment of neonatal aortic coarctation (A) Balloon angioplasty samples (B) Stented samples (i) Haematoxylin and Eosin (H&E) (ii) Picrosirius red (PS-red) (iii) Polarized light microscopy (iv) Verhoeff elastin (VE). Scale bars (Black = 1mm, White = 250 µm)

Looking at the stented samples, locations of high ductal tissue volume appear to have permanent stent indentations whilst the more aortic type regions show more recoil in comparison, see figure 6. This further supports observations made in Johnston and Linnane et al (2024) [Johnston &Linnane 2024] that due to the microstructural composition of the ductal tissue, it is more compressible by nature.

**Figure 6:**
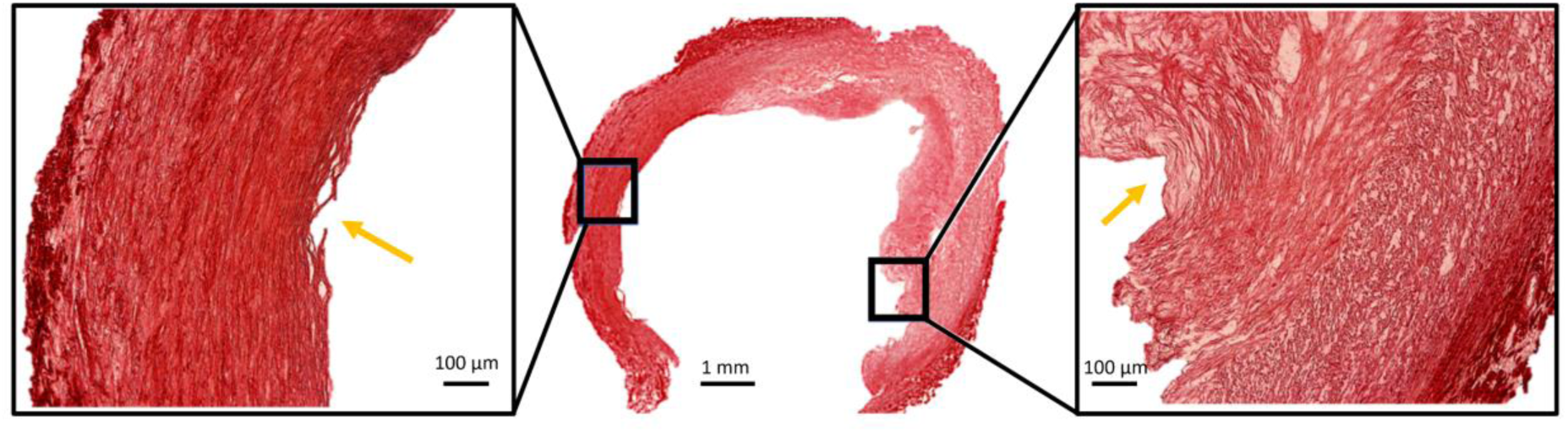
Stented neonatal coarctation sample 8 showing more pronounced indentation of regions of ductal tissue (right side) in comparison to the healthier aortic component (left side). Yellow arrows highlight area of stent indentation.

### Collagen Hybridizing Peptide (F-CHP) of ballooned and stented aortic coarctation samples

Firstly, it must be stated that there is some background fluorescence noted in our F-CHP-stained controls and this can be because of two reasons, 1) there is a low content of degraded collagen content present or 2) there is autofluorescence from the tissue itself. F-CHP quantification shown in figure 7 demonstrates that there is an increase in the F-CHP fluorescence when comparing the ballooned neonatal aortic coarctation samples to the respective control group. Statistical analysis in figure 7B shows the mean fluorescence is significantly different in control samples compared to ballooned samples, suggesting collagen denaturation occurring within the tissue due to balloon expansion. To understand why this mean fluorescence is higher in the ballooned group compared to the control group, the isthmus balloon ratio was used to group samples and investigate the stretch imposed on the tissue during balloon expansion. As shown by figure 7C, it is noted that samples that are stretched further (>2) due to balloon inflation show a significantly higher mean F-CHP fluorescence than less stretched samples (<2), demonstrating that there is increased collagen denaturation at higher strains.

**Figure 7:**
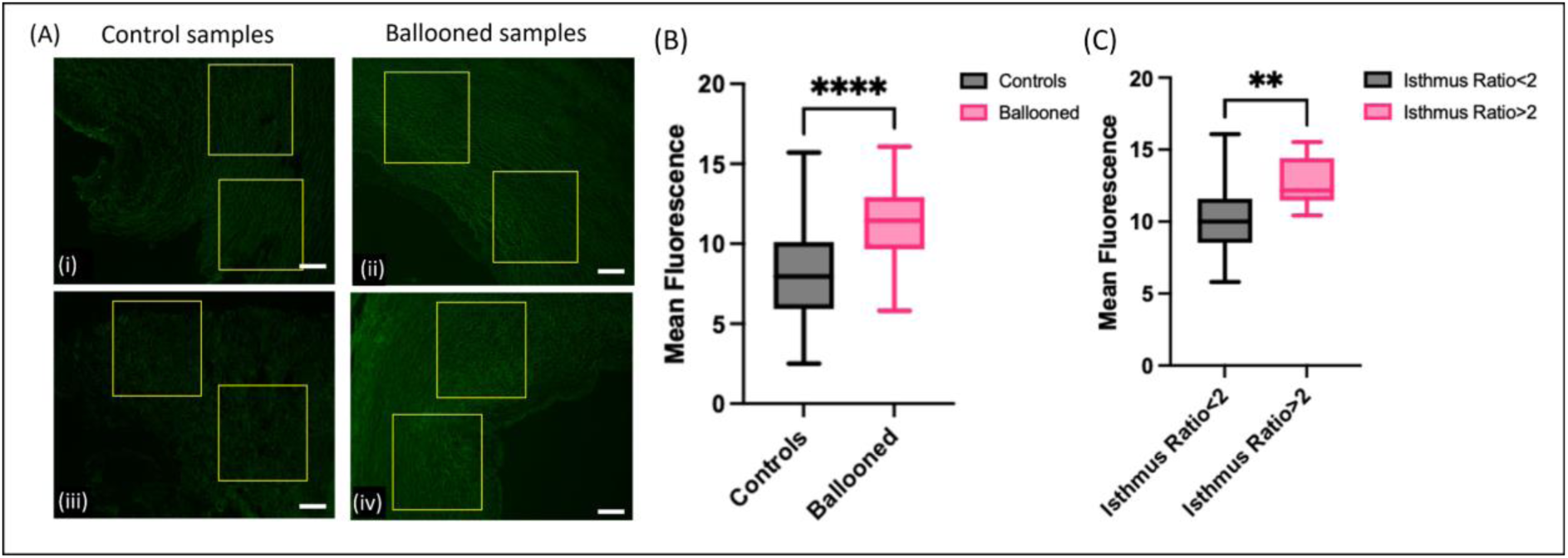
(A) F-CHP images of controls ((i) and (iii)) and representative ballooned samples ((ii) and (iv)) showing visible increase in the fluorescence intensity when samples have been ballooned. Scale bar = 50 µm (B) Unpaired t-test showing significant difference in the mean intensity between control and ballooned samples (P = 0.001) (C) Mean fluorescence observed in ballooned samples when sorted into grouping with isthmus/balloon ratio greater than 2 and less than 2.

Visually, the F-CHP fluorescence in our stented neonatal aortic coarctation samples looks significantly brighter, see figure 8A, than the controls and balloon expanded neonatal aortic coarctation samples shown in figure 6A. A major difference is that there is clear depth varying stent strut indentations (yellow arrows) on the tissue when compared to the ballooned samples, supporting the histological observation of the variable microstructural composition. Like the ballooned neonatal aortic coarctation samples, there is a statistically significant increase in the mean fluorescence in the stented samples versus the control samples, see figure 8B. Interestingly, when the level of stretch was analysed by grouping with respect to the balloon to isthmus ratio and grouped with ballooned samples, the mean fluorescence was significantly increased when compared to the controls, see figure 8B. Similar to the balloon angioplasty result, due to the tissue experiencing higher strain due to stent expansion, it is observed there is higher CHP fluorescence suggesting further denaturation of the collagen during this procedure, see figure 8C.

**Figure 8:**
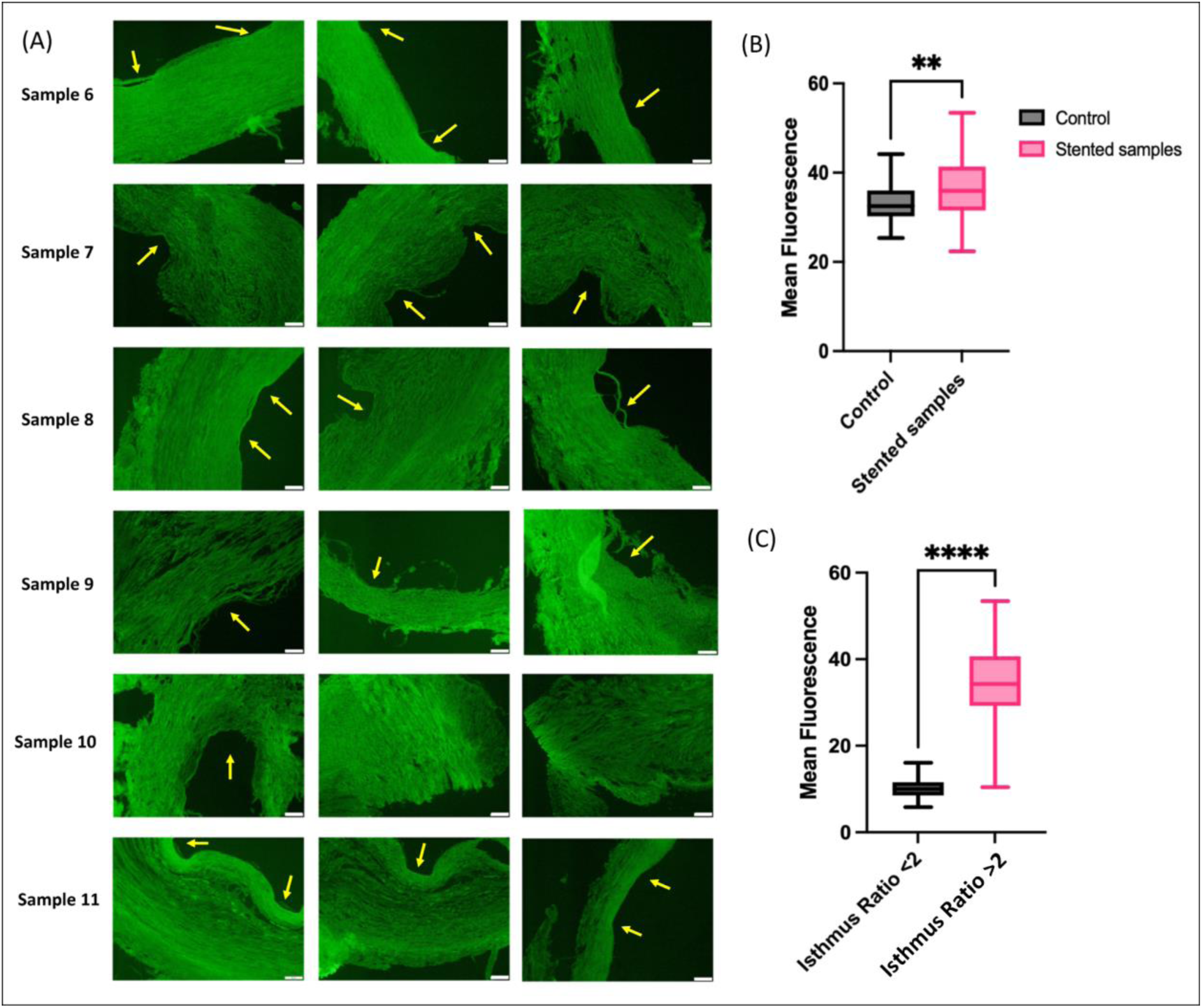
(A) F-CHP images of stented neonatal aortic coarctation samples with noticeable stent indentations (yellow lines). Noticeable variation of fluorescence throughout the tissue. Scale bar = 50 µm (B) Unpaired t-test showing significant difference between mean fluorescence of control and stented neonatal aortic coarctation (C) Mean fluorescence observed in ballooned and stented neonatal aortic coarctation samples when sorted into grouping with isthmus/balloon ratio greater than 2 and less than 2 (P < 0.001).

Like the results observed in figure 6, it is clearly observed with locations of high ductal tissue volume that the stent permanently indents the sample whilst the healthier region shows more recoil in comparison, see figure 9. An interesting observation is the variability in fluorescence observed within the sample in which the healthier side shows a higher intensity (red arrows) on the more luminal side with little indentation (yellow arrows), shown by figure 9A. Meanwhile on the higher ductal volume side it is observed there is significant indentation (yellow arrows) of the stent within the tissue, with higher fluorescence then observed in the healthier tissue present through the thickness (red arrows), see figure 9B. This observation supports previous assessments that due to microstructural composition of the ductal tissue; it is compressible by nature with little to no collagen or elastin present. Furthermore, due to the lack of collagen in particular, the F-CHP intensity is lower in the ductal tissue when compared to healthy tissue.

**Figure 9:**
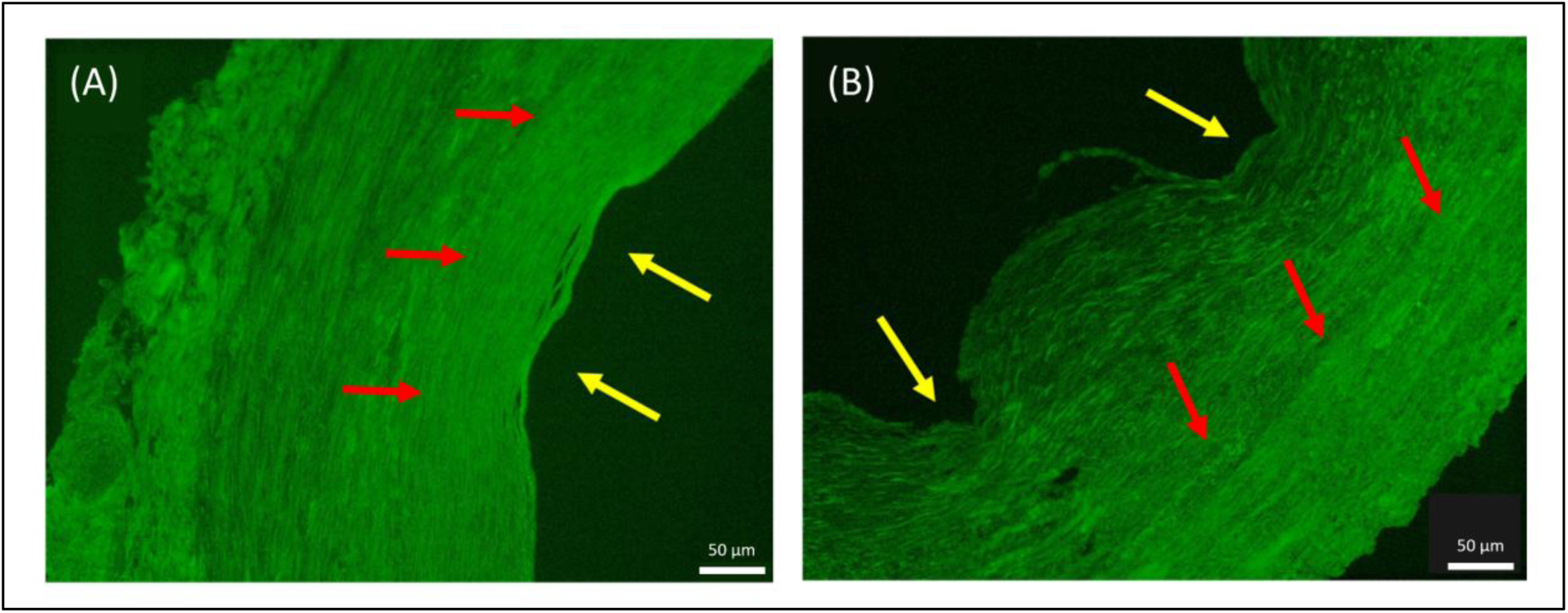
F-CHP of stented neonatal aortic coarctation sample 8 showing more pronounced indentation of region of ductal tissue (right side) in comparison to the healthier aortic component (left side). Yellow arrows highlight area of stent indentation with red arrows highlighting the areas of higher fluorescence in healthier regions of the tissue.

An advantage of using F-CHP in quantification is that it allows for regional assessment of the level of collagen damage to be established within the tissue. As shown histologically by figure 10A, there is clear microstructural variation throughout neonatal aortic coarctation samples. Not only is there a variance in the elastin content as shown in previous publications, but there is also variability in the collagen content. When the sample is stented, F-CHP can identify this variable structure with the bonus of being able to quantify specific regions within the tissue subjected to higher levels of stretch and therefore collagen damage. As shown by figure 10B and the histograms, low intensity fluorescence distribution is observed at the highly indented regions (yellow arrow) which histologically shows an initial ductal rich presence. An increased fluorescence intensity distribution is then observed once the region of interest is in the healthier load bearing region of the tissue, which is collagen and elastin rich. Importantly, this highlights 1) the compressible nature and low damage occurring within ductal parts of the tissue during stent expansion and 2) the load bearing healthier tissue experiencing higher damage to support the outward load imposed by stenting.

**Figure 10:**
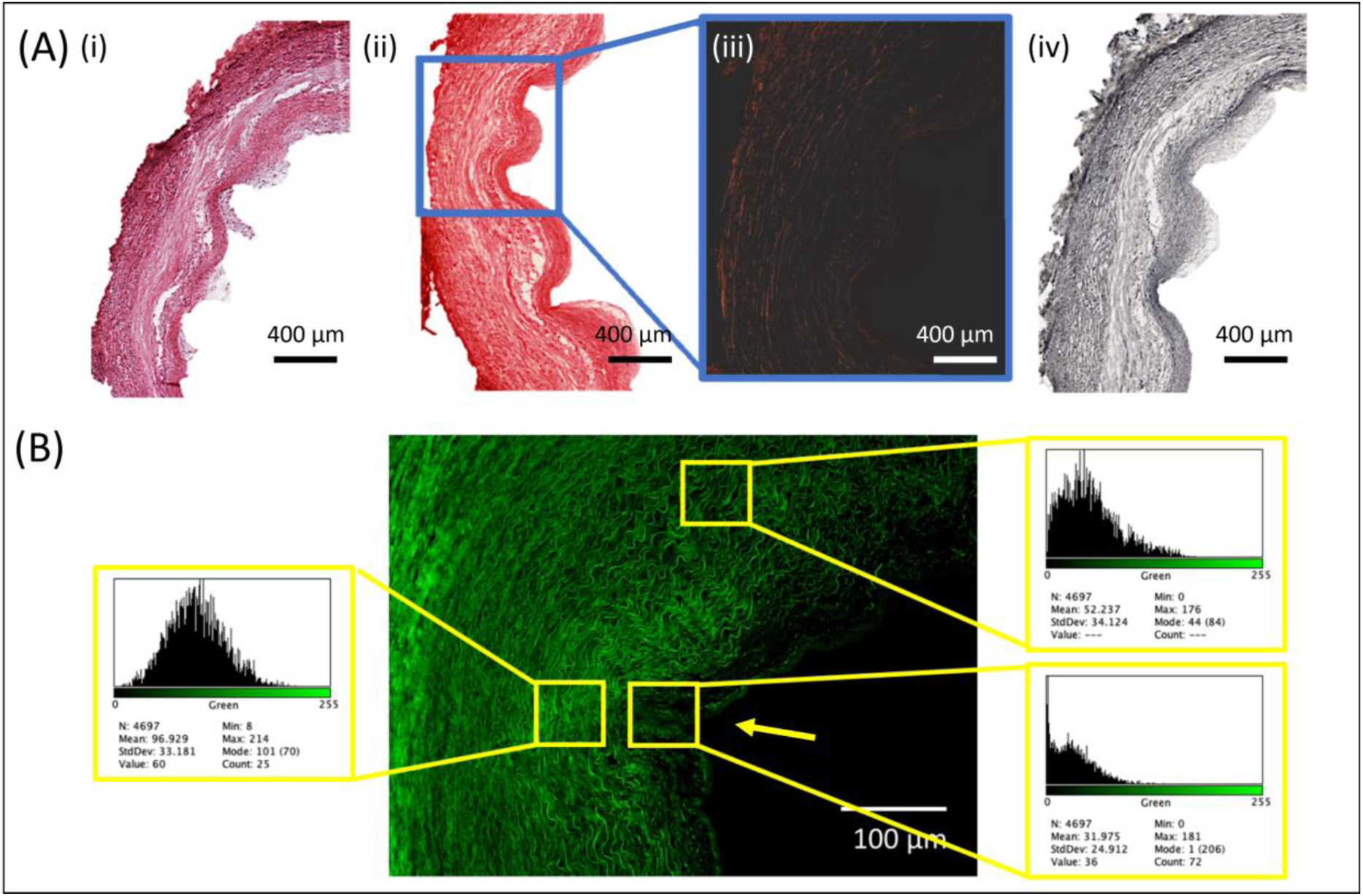
Regional specific assessment of F-CHP fluorescence in stented neonatal aortic coarctation samples (A) Histological overview with (i) H&E, (ii) PS red, (iii) PLM and (iv) VE (B) Regional assessment with fluorescent intensity histograms showing varied distribution throughout the sample and highly localized damage in regions.

## Discussion

It has been well described that the predominant feature of neonatal aortic coarctation from histological assessment is the lack of elastic laminae [54–57]. This is thought to play a role in the limited capability for the tissue to undergo stretch in response to an intervention. Our histological results presented for ballooned and stented native aortic coarctations agree with those previous studies and our previous study [Johnston &Linnane 2024], where it is clearly demonstrated that there is a non-consistent and reduced number of elastic laminae in the ductus arteriosus part of the neonatal aortic coarctation specimens. It has been established that collagen is a key load bearing constituent in arterial vessels [58–62] and it is important to note that from this work there is a paucity of collagen present within the ductal arteriosus tissue in CoAs. Overall, there is a striking difference between the ductal arteriosus tissue and the surrounding aortic tissue, showing a healthier microstructure which is both collagen and elastin rich. Whether this healthier tissue is comparable in terms of content to healthy aortic tissue is still unknown. Regardless, at the coarctation site where the lumen of the vessel was narrowest, the cause of the narrowing was due to ductal arteriosus tissue being present.

The first aim of this study was to investigate the neonatal coarctation response to percutaneous treatment options such as balloon angioplasty and/or stent insertion. It has been noted in the literature that balloon angioplasty has largely gone out of favour since the advent of greater stent options due to the need to create intimal and medial tears to achieve lumen gain [63,64]. As outlined in the histological results, the ductus arteriosus tissue seen in the CoA specimens has little elastin or collagen and thus very little structural integrity. This was clearly demonstrated in the balloon angioplasty experiments where ultrasound images show the balloon clearly and easily compresses the ductal tissue but once deflated the lumen closes over again. Thus, inferring that the ductal tissue cannot be deformed and without causing deformation to the aortic tissue, sufficient lumen gain will not be achieved. With a larger balloon (6mm) and where the aortic tissue clearly undergoes more stretch, there is more obvious deformation, and the lumen remains patent even after balloon deflation (4.7mm to 6.7mm), even though there is still significant tissue recoil. While these experiments are limited by the lack of internal blood pressure which arguably may have maintained some lumen gain, it is observed that unless the tissue is stretched well beyond supraphysiological levels, little lumen gain will be achieved by angioplasty alone, justifying the move to performing stenting of the CoA. When stenting, it is observed in 5 out of the 6 cases, that the Minima opened the vessel to the desired lumen diameter which verifies observations made in a previous animal study that the target lumen diameter can be achieved [26]. Furthermore, no stent failure was observed in all cases, demonstrating the Minima stent’s structural integrity and the feasibility of being implanted within neonatal aortic coarctations. It was observed from our histological results that sample 10, which failed during stenting, had high ductal tissue volume when compared to other samples and that failure occurred at the site where the ductal tissue propagated through the entire thickness of the tissue. This is an interesting observation and suggests a different stenting approach should be utilized to ensure that failure does not occur. A recent study by Cornelissen et al (2023) has demonstrated that the level of stent indentation can be linked to tissue damage [30]. The authors used optical coherence tomography (OCT) to calculate the level of indentation of the stent struts and linked this to histological damage scores. This application is very interesting and could be adapted to be used in stenting procedures. As demonstrated by the results in the current study and our previous study [Johnston &Linnane 2024], the ductal tissue is readily compressible and easily indents with the implantation of a stent. OCT can be used to assess how much ductal tissue is present in the coarctation region during a stenting procedure and subsequently stents can be serially expanded to limit the level that healthy aortic tissue is indented, and therefore limit damage induced to the vessel.

The second aim of this study was to investigate and quantify the level of microstructural damage in neonatal aortic coarctation when undergoing balloon angioplasty and stenting. Collagen hybridising peptide (CHP) is a peptide which has been designed to bind to damaged collagen and thus enable a quantification of collagen damage using fluorescence [40]. Furthermore, CHP has been used as a biomarker for early onset atherosclerosis [34]. The results clearly demonstrate an increase in the level of fluorescence between tested samples (ballooned and stented) and controls which indicate the denaturation of collagen during these procedures. Furthermore, when comparing the stenting to the ballooned cases, increased vessel stretches induced higher denaturation of the collagen and therefore higher fluorescent intensities are observed. This increased fluorescence with increased stretching of the tissue agrees with previous studies such as Converse et al (2017), Zitnay et al (2017) and Anderl et al (2023) [35,37,41]. The interesting observation from this work is due to the varying composition of the tissue itself where varying fluorescent intensities are present. Furthermore, in regions of high stent strut indentation, there is a decreased fluorescent intensity when compared to regions with low stent strut indentation depicting a higher intensity. Our histological results can explain that this high stent strut indentation and low fluorescent intensity combination is due to the low collagen & elastin content within the compressible ductal tissue, with the opposite being observed for “healthier” regions of the tissue and this is highly important. This allows CHP to be used not only as a metric for collagen denaturation, as it has previously and in this study, but ensures that variable microstructural compositions within the tissue can be identified along with how much it has been damaged before treatment. This paves the way for patient specific stents to be designed and developed, to achieve the desired lumen diameter and reduce vessel damage. Lastly, despite the limitations associated with using CHP, increased levels of stretch do lead to higher levels of fluorescence and therefore, more collagen damage. This indicates that the greater the stretch the tissue undergoes, the more collagen that is damaged and possibly the greater the aneurysm risk in the long term. This fits with observations in the literature whereby balloon angioplasty has a higher aneurysm rate than surgery or stent placement [63], likely as intimal and medial damage must occur during balloon angioplasty to achieve the required lumen gain. Since lumen gain is the goal of the percutaneous strategies, the CHP results indicate that direct inflation to the maximum diameter with stent deployment will induce collagen damage during such procedures. Therefore, a staged approach to stenting may be necessary for optimum outcomes such that by minimising the stretch at each stage of stent deployment the amount of initial collagen damage may be reduced, allowing the tissue to remodel to accommodate this change to reduce the aneurysm risk.

There are some limitations to this study that should be mentioned. Firstly, the neonatal aortic coarctation tissue that underwent both balloon angioplasty and stenting was in an unloaded configuration, with no inclusion of blood pressure. The benchtop setup could be amended to incorporate a pump system to replicate this blood pressure change in the vessel and is of interest in future studies to assess damage accumulation over time. Secondly, recent studies have been looking at patient specific stents for the possible treatment of CoA [65] whilst the stent used in this study is the Minima stent and is not patient specific. Future work aims to manufacture bespoke stent designs using 3D printing and an optimization approach to minimize the vessel stress whilst achieving the target lumen diameter. Lastly, stents were expanded straight to 6mm for our stenting experiment with the same stent used in each test. Future work will look at different deployment strategies in combination with different stent designs to establish the best combination that achieves the desired lumen diameter whilst reducing the level of tissue damage during the procedure.

## Conclusions

Neonatal aortic coarctations is an extremely heterogenous disease with tissue samples demonstrating highly variable microstructural composition. Performing a balloon angioplasty procedure on its own is not sufficient to achieve the desired lumen gain, with tissue recoil causing collapse of the lumen diameter once the balloon is deflated. The Minima stent can be successfully deployed to stent neonatal aortic coarctation tissue and achieve the desired lumen diameter, although there is increased collagen damage observed in doing this. This work suggests that future stenting procedures should consider the tissue’s microstructural composition before treatment and implement an incremental deployment strategy to optimize stent performance and reduce microstructural damage.

## Data Availability

All data produced in the present study are available upon reasonable request to the authors

## Ethical Approval

Ethical approval for obtaining the samples and imaging used in this study was obtained from the research and ethics board of Children’s Health Ireland. Informed consent was obtained from the parents / legal guardians of all patients for the tissue samples used in this study.

## Acknowledgements

The authors would like to thank Dr Evan Zahn, Eason Abbott and Dustin Armer from Renata Medical for providing the stents used in this study. The authors would like to acknowledge the Cardiothoracic Surgical team, especially, Mr Jonathan McGuinness, Prof. Lars Nolke and Prof. Mark Redmond, in Children’s Health Ireland at Crumlin for aiding in obtaining the neonatal coarctation samples for tissue testing and CHP analysis.

